# Adverse health impacts of cooking with kerosene: A multi-country analysis within the Prospective Urban and Rural Epidemiology Study

**DOI:** 10.1101/2020.06.19.20135871

**Authors:** Raphael E Arku, Michael Brauer, MyLinh Duong, Li Wei, Bo Hu, TSE Lap Ah, Prem K Mony, PVM Lakshmi, Rajamohanan K Pillai, Viswanathan Mohan, Karen Yeates, Lanthe Kruger, Sumathy Rangarajan, Teo Koon, Salim Yusuf, Perry Hystad, PURE (Prospective Urban and Rural Epidemiological) Study investigators

**Author notes:** Correspondence to: Raphael E Arku, School of Public Health and Health Sciences, University of Massachusetts, Amherst, MA, USA.

## Abstract

**Background:** Kerosene, which was until recently considered a relatively clean household fuel, is still widely used in low and middle-income countries for cooking and lighting. However, there is little data on its health effects. We examined cardiorespiratory effects and mortality in households using kerosene as their primary cooking fuel within the Prospective Urban Rural Epidemiology (PURE) study.

**Methods:** We analyzed baseline and follow-up data on 31,490 individuals from 154 communities in China, India, South Africa, and Tanzania where there was at least 10% kerosene use for cooking at baseline. Baseline comorbidities and health outcomes during follow-up (median 9.4 years) were compared between households with kerosene versus clean (gas or electricity) or solid fuel (biomass and coal) use for cooking. Multi-level marginal regression models adjusted for individual, household, and community level covariates.

**Results:** Higher rates of prevalent respiratory symptoms (e.g. 34% [95% CI:15-57%] more dyspnea with usual activity, 44% [95% CI: 21-72%] more chronic cough or sputum) and lower lung function (differences in FEV_1_: -46.3 ml (95% CI: -80.5; -12.1) and FVC: -54.7 ml (95% CI: -93.6; -15.8)) were observed at baseline for kerosene compared to clean fuel users. The odds of hypertension was slightly elevated but no associations were observed for blood pressure. Prospectively, kerosene was associated with elevated risks of all-cause (HR: 1.32 (95% CI: 1.14-1.53)) and cardiovascular (HR: 1.34 (95% CI: 1.00-1.80)) mortality, as well as major fatal and incident non-fatal cardiovascular (HR: 1.34 (95% CI: 1.08-1.66)) and respiratory (HR: 1.55 (95% CI: 0.98-2.43)) diseases, compared to clean fuel use. Further, compared to solid fuel users, those using kerosene had 20 – 47% higher risks for the above outcomes.

**Conclusions:** Kerosene use for cooking was associated with higher rates of baseline respiratory morbidity and increased risk of mortality and cardiorespiratory outcomes during follow-up when compared to either clean or solid fuels. Replacing kerosene with cleaner-burning fuels for cooking is recommended.

**Highlights:**

- Kerosene was (until 2014) considered a relatively clean household fuel for cooking and lighting
- Though the WHO discouraged kerosene use, evidence in support of this position are still scarce
- We prospectively examined the effects of kerosene use on cardiorespiratory morbidity and mortality
- Kerosene use for cooking was associated with higher rates of baseline respiratory morbidity
- Prospectively, kerosene use was associated with increased risk of mortality and incident cardiorespiratory outcomes compared to clean fuel or solid fuel use
- Replacing kerosene with cleaner-burning fuels for cooking is recommended

## Introduction

Kerosene, also known as paraffin or lamp fuel, is an important household fuel that continues to be widely used in developing countries by almost a billion people for cooking and lighting (Lam et al. 2012b; Mills 2005; Mills 2017). As a fractional distillate of petroleum oil with a mixture of short-chain hydrocarbons, kerosene is also used in several commercial (e.g. aviation fuel) and industrial (e.g. general solvents) applications. Cooking primarily with kerosene is predominant in African and South-East Asian regions (World Health Organization 2016). For example estimates suggest that 80% of households in Djibouti, and between 25-34% in Equatorial Guinea, Eritrea and Nigeria use kerosene as their primary cooking fuel (World Health Organization 2016). Until 2014, “primary reliance on solid fuels for cooking”, which serves nearly 3 billion people globally (over 75% in sub-Saharan Africa), was the indicator for dirty fuels (World Health Organization 2014). As a result, kerosene has been regarded among the “cleaner-burning” household fuels in comparison with solid fuels (biomass and coal), and has been grouped with liquefied petroleum gas (LPG), biogas, ethanol, and electricity as a “modern” fuel (Lam et al. 2012b; Legros et al. 2009; World Health Organization 2014). Besides the focus on “solid” fuels, the designation of kerosene as a “clean” fuel was in part due to views of some international agencies and academic researchers which considered kerosene as less polluting in the larger context of household fuels (Grieshop et al. 2011), and was being subsidized for affordability (Melsom et al. 2001; Mills 2017; UNDP/ESMAP 2003). While a few studies of kitchen area and personal exposure levels in homes cooking with kerosene have documented high fine particulate matter (PM_2.5_) levels (Andresen et al. 2005), emissions estimates for common pollutants from kerosene combustion, including fine particles, are close to those of LPG and well below those of charcoal, which has been characterized as a moderately polluting fuel (Grieshop et al. 2011). As a result, until 2014 when kerosene was declared a polluting fuel and its use as a household fuel discouraged (World Health Organization 2014), the exposures to and risks associated with the use of kerosene for cooking have not been well-documented nor received the attention given to solid fuels (Lam et al. 2012a).

In experimental studies, kerosene combustion has been found to result in diverse biological responses including inflammation, production of reactive oxygen species, hypoxia, immunosuppression, reduced effectiveness of pulmonary surfactants, and alterations in membrane lipids (Lam et al. 2012b; Maiyoh et al. 2015). Emerging epidemiologic studies from several low- and middle-income countries (LMIC) have also shown significant adverse effects associated with kerosene use for cooking (World Health Organization 2014), including elevated blood pressure (BP) (Alexander et al. 2017; Arku et al. 2017); adverse pregnancy outcomes (Alexander et al. 2018; Andresen et al. 2005); impaired lung function and respiratory illnesses (Bates et al. 2018; Bueso et al. 2010; Coker et al. 2020; Dagoye et al. 2004; Elf et al. 2019; Lam et al. 2012b; Pokhrel et al. 2010; Ranathunga et al. 2019; Venn et al. 2001); cancers, and deaths (Lam et al. 2012b; Mitter et al. 2016). However, these studies were small in sample sizes and limited in geographical coverage. Although kerosene is still widely used for cooking in an estimated 200 million households in LMICs (World Health Organization 2016), and despite the current shift in global understanding that kerosene is no longer considered a ‘clean’ fuel for health, there has been a lack of relevant health effects studies of cooking with kerosene in LMICs (Lam et al. 2012b). Specifically, there are no prospective cohort studies of the adverse health impacts of cooking with kerosene. The prospective study design enabled us to identify and follow a large population of kerosene users over time in order to provide strong scientific evidence about the link between kerosene use and several outcomes of interest simultaneously, including incident disease, while reducing selection and recall bias.

We therefore leverage a large ongoing international cohort – the Prospective Urban Rural Epidemiology (PURE) Study – to evaluate the effects of primary use of kerosene for cooking, compared to clean (electricity or LPG), and solid fuels, on a range of health measures. We examined the association between baseline cardiorespiratory morbidity (respiratory symptoms, lung function, BP and hypertension), and prospectively documented mortality, and incidence of major cardiovascular (CVD) and respiratory disease during follow-up (median of 9.4 years) in 31,490 individuals from 154 communities in China, India, South Africa, and Tanzania. With one of the largest samples and geographical coverage, our findings fill an important knowledge gap and add to the limited data currently available on the impact of cooking with kerosene on cardiorespiratory health. We also provide new data on the effects of kerosene use relative to solid fuels.

## Methods

### Study design and participants

PURE is a community-based cohort of ∼165,000 participants aged 35–70 years living in 750 urban and rural communities in 21 low, middle and high-income countries (Teo et al. 2009; Yusuf et al. 2020) (Figure 1). The communities represent neighborhoods in urban areas and small villages in rural areas, aggregated into study centers according to their geographical cluster. Harmonized approaches were used to enumerate households and to recruit individual participants, with baseline measurements in this analysis conducted between 2001-2011 and follow-up between 2002-2018. Trained personnel collected individual, household, and community-level information at baseline. Individual level information on demographic factors, lifestyle behaviors, medical history, spirometry, BP, anthropometrics, and blood samples were collected. At the household level, data on primary cooking fuels were gathered.

**Figure 1.**
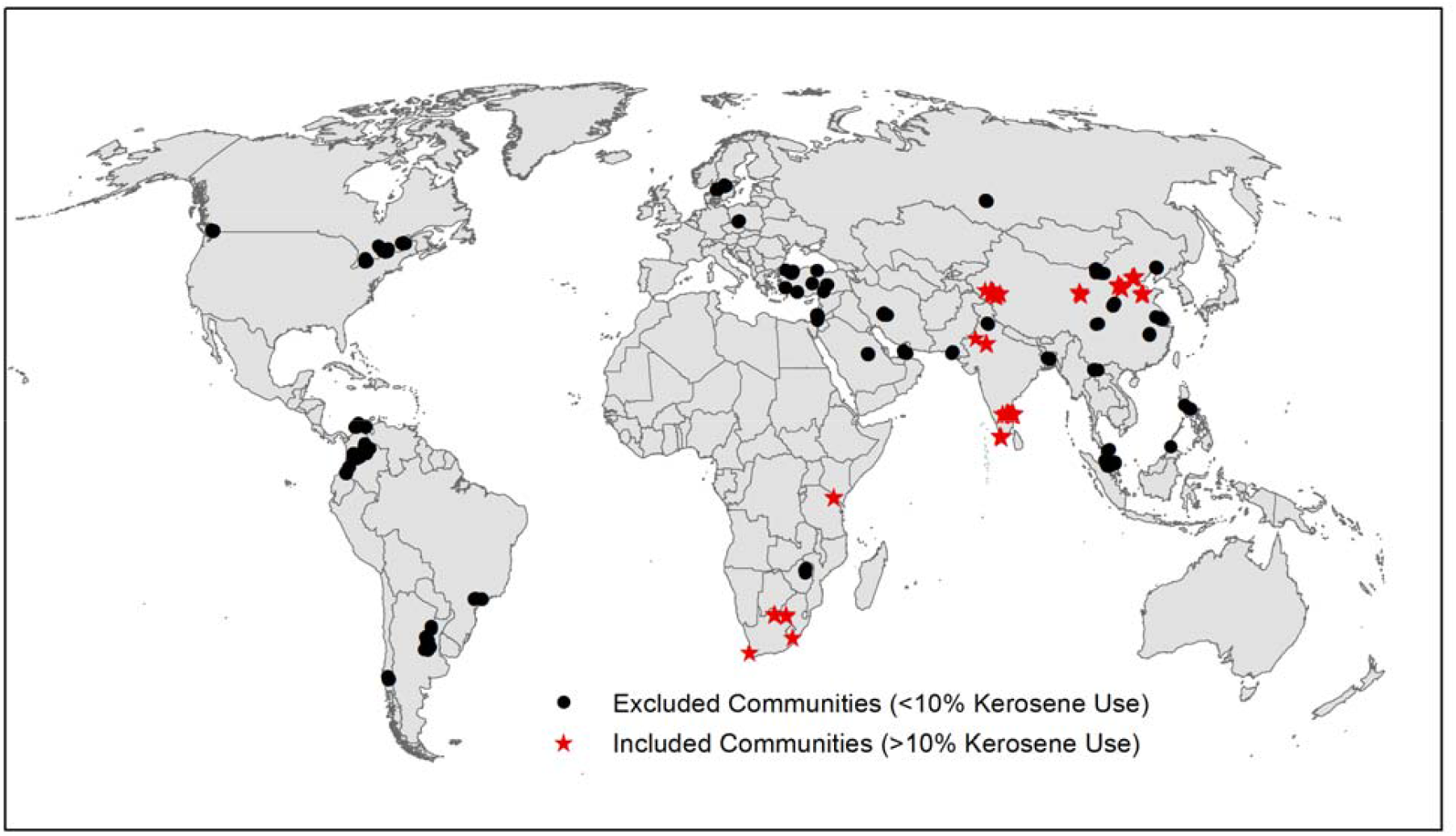
**Map of PURE communities included in this analysis. Communities were included if they were located in centers that reported ≥10% of primary use of kerosene for cooking at baseline**.

This analysis of primary use of kerosene for cooking included all participants in centers in China (five centers); India (four centers); South Africa (two centers); and Tanzania (one center) where there was at least 10% kerosene use at baseline (Figure 1). The criterion to include only study centers with at least 10% kerosene use was decided a priori to ensure sufficient numbers of kerosene users to enable valid comparisons between the exposed and unexposed groups. In each center, we recorded each household’s primary cooking fuel, coded as: kerosene, LPG, biogas, electricity, or solid fuel (wood, coal, charcoal, agricultural products, shrub/grass, and animal dung). However, the data did not include information on secondary fuels or stove/fuel stacking (i.e. whether there was parallel use of different fuels and technologies in the same home). For the centers included in this analysis, baseline data collection and follow-up periods ranged 2001-2018 (India), 2005-2018 (China), 2005-2018 (South Africa), and 2011-2018 (Tanzania).

The PURE study (and this analysis) was approved by the Institutional Review Boards of Hamilton Health Sciences, Oregon State University, the University of British Columbia (H14-02982), and the local ethics committees in the participating countries.

### Outcomes of interest

We included participants with complete data on exposure (i.e. household’s primary use of kerosene for cooking) and at least one outcome of interest. Thus, the sample sizes for analysis varied by individual outcomes as detailed below. We examined self-reported cardiorespiratory symptoms in the six months preceding study enrollment, and BP and spirometry measured at baseline. We also examined follow-up data on all-cause and cause-specific mortality, and incidence of major cardiorespiratory events.

#### Baseline data

Questionnaires were used to gather information on baseline respiratory symptoms and co-morbidities occurring at least weekly in the previous six months: self-reported wheeze; cough or sputum; dyspnea; and chest pain or tightness with usual activity.

Blood pressure was measured during the morning in sitting position using Omron digital BP measuring devices (Omron HEM-757) (Arku et al. 2020; Chow et al. 2013; Yusuf et al. 2004). Trained personnel took two systolic and diastolic BP measurements from the right arm about one minute apart using standardized procedures. Participants refrained from smoking, ingesting food or caffeine beverages, and exercising (including stair climbing) 30 minutes prior and further had to sit quietly for 5 minutes prior to all clinical measurements. The mean of the two BP measurements were recorded. Hypertension was defined as an average systolic BP ≥ 140 mm Hg, an average diastolic BP ≥ 90 mm Hg, or self-reported hypertension with maintenance anti-hypertensive therapy.

Spirometry was conducted following the American Thoracic Society guidelines (Miller et al. 2005) using a portable MicroGP Spirometer (MicroMedical, Chatham IL, USA). Calibration was performed at least monthly or prior to each use. Up to six maneuvers were attempted by each participant while standing and wearing a nose clip. The maneuvers were performed with maximal effort and observed for forced exhalation to six seconds or longer without coughing. The two highest measurements of forced expiratory volume in 1 second (FEV1) and forced vital capacity (FVC) within 200 mL variability were recorded. The highest FEV1 and FVC were analyzed. In addition to analyzing FEV1 and FVC as continuous measures, we used the FEV1/FVC ratio <0.70 cutoff as an indication of airflow obstruction, and coded as (yes/no) (Jing et al. 2009; Rabe et al. 2007) for further analysis. The spirometry data in PURE have previously been evaluated for external, internal, and face validity (Duong et al. 2013).

#### Follow-up data and events

We included follow-up data on all-cause mortality, major cardiovascular (any CVD death and incidence of non-fatal myocardial infarction, stroke, and heart failure), and major respiratory (any respiratory death and non-fatal tuberculosis, chronic obstructive pulmonary disease, pneumonia and lung cancer) disease. Participants were contacted at least once every three years to document events and diseases. Due to the limited coverage of vital registration systems in several LMIC in PURE, event identification relied on combinations of face to face interviews with the participants or their household members, certified hospital records, death certificates, and verbal autopsies where necessary (Gajalakshmi et al. 2002), to assess the cause of deaths or hospitalization. Following a standardized protocol, trained physicians also used several sources of information on CVD events and all deaths to adjudicate outcomes in each county. In addition, a random sample of events from each center was assessed at the study coordinating center for further adjudication to ensure internal consistency (Dehghan et al. 2017; Teo et al. 2009; Yusuf et al. 2014).

### Statistical analysis and covariates

We estimated associations of self-reported primary use of kerosene for cooking relative to clean (gas or electricity) fuel using multilevel marginal regression models. Logistic regression was used to evaluate the odds of having chronic wheeze, cough or sputum, dyspnea with usual activity, chest pain or tightness with usual activity, elevated BP (hypertension), and the FEV1/FVC ratio <0.70 in kerosene users relative to the use of clean fuels at baseline. We used linear regression to compare the cross-sectional averages in baseline systolic and diastolic BP, FEV1, FVC, and FEV1/FVC ratio, among kerosene versus clean fuel users. To further assess whether there were relatively larger decreases in lung function across age groups of kerosene relative to clean fuel users, we compared the cross-sectional age-related differences in baseline FEV1/FVC ratio for kerosene use relative to clean fuels. In longitudinal analysis, Cox proportional hazards models were used to estimate the occurrences and hazard ratios (HRs) for all-cause mortality, and major cardiorespiratory events and deaths, among kerosene users compared to clean fuel users. Finally, to evaluate the overall major adverse impact of primary use of kerosene for cooking, we combined all first major CVD and respiratory disease, or death into a composite outcome using an indicator variable; this composite measure was evaluated separately in a Cox proportional hazards model.

Models were adjusted for different sets of covariates depending on relevance to the specific outcome measure, with relevant covariates including age (continuous), sex (male/female), education (≤ primary, secondary, higher), body mass index (BMI; continuous), smoking (current, former, never), second hand smoke exposure (yes/no), alcohol use (current, former, never), rural-urban location, wealth index (an indicator of household wealth generated based on household’s owned items standardized by tertiles for each country), physical activity (assessed as low, moderate, high according to the International Physical Activity Questionnaire (Craig et al. 2003)), INTERHEART risk score (a validated score indicative of cardiovascular risk-factor burden (McGorrian et al. 2011)), occupational class (missing, homemaker, unskilled, skilled, professional), proportion of household income spent on food (percent), and baseline long-term average outdoor fine particulate matter pollution (PM_2.5_; particles below 2.5 micrometers in aerodynamic diameter) derived from a global geographically weighted regression model using data from various satellite-, simulation-, and ground monitor-based sources (van Donkelaar et al. 2016). PM_2.5_ concentrations were estimated at 1 × 1 km resolution for years before, at, and after study enrollment: 2001 – 2011. Community long-term average PM_2.5_ concentrations were defined as the 3-year average centered on the year of enrollment (i.e. average of the annual estimates at the year before, at, and after enrollment; see Arku et al. (Arku et al. 2020) for additional details). We evaluated two models for each outcome. Minimally adjusted models accounted for age, sex, country, center and rural-urban status. (Cox proportional hazards models included a strata variable for center). Final models additionally adjusted for behavioral and clinical covariates as well as socioeconomic indicators. As very few households (< 20%) had multiple participants, adding a household random effect along with country and center random effects in the models did not change the results. Therefore, we did not include in the models a random effect factor for household.

In further analyses to evaluate potential residual confounding, we first conducted the analyses without the following poverty related variables and compared the results with the full model: education, wealth index, occupation, % income spent on food (classified as ≥ 50% or < 50%, an indication of medium food security (Smith and Wiesmann 2007)). Next, we stratified models by the above poverty related variables identified a priori along with urban/rural and country as these variables could modify the impact of kerosene use on measured outcomes. Further, using the exact same models for kerosene and clean fuel users, we compared the risk associated with primary use of kerosene for cooking to primary use of solid fuel for all outcomes. This was done for two reasons: to assess the relative effect of kerosene versus solid fuels for all outcomes; and to evaluate potential for residual confounding by sociodemographic factors as they were likely to be more similar between kerosene and solid fuel users than between kerosene and clean fuels users.

Analyses were conducted in the open-source statistical package R version 3.4.1 (R Project for Statistical Computing, 2016) and SAS version 9.4.

## Results

We included 31,490 participants between the ages 35-70 years from 154 communities at 12 centers in China, India, South Africa, and Tanzania (Figure 1 and Table 1). The proportion of households using kerosene was ∼10% in both urban (n=23,219) and rural (n=8,271) areas (Table 1). The mean age was similar between kerosene and clean fuel users (∼ 50 years). Compared to those using clean fuels, kerosene users were more often female, and had lower measures of SES, including lower education and wealth index, and were more likely to engage in unskilled work.

**Table 1.** **Characteristics of 31**,**490 individuals living in centers with >10% kerosene use for cooking at baseline, stratified by urban-rural status and kerosene versus clean fuels (electricity or gas) as primary cooking fuel**

| Variables | Urban (n=23,219) |  | Rural (n=8,271) |  |
| --- | --- | --- | --- | --- |
|  | Kerosene | Clean fuel | Kerosene | Clean fuel |
| No. Individuals | 2025 | 21194 | 742 | 7529 |
| No. Communities | 44 | 69 | 52 | 87 |
| Female (%) | 65 | 57 | 63 | 59 |
| INTERHEART risk Score<sup>a</sup> ($\bar{x}$, $sd$) | 8 (5) | 9 (5) | 8 (5) | 9 (5) |
| Low education (≤ Primary School) (%) | 73 | 20 | 72 | 42 |
| Unskilled worker (%) | 57 | 23 | 69 | 72 |
| % of Income spent on Food ($\bar{x}$, $sd$) | 57 (27) | 43 (21) | 49 (30) | 45 (26) |
| Lowest tertile home wealth index (%) | 31 | 6 | 57 | 30 |
| Ever Smoker (%) | 36 | 23 | 30 | 27 |
| Regular second-hand smoke exposure (%) | 16 | 31 | 32 | 22 |
| Alcohol Use (former & current) (%) | 34 | 20 | 26 | 22 |
| BMI ($\bar{x}$, $sd$) | 24 (6) | 25 (4) | 25 (6) | 25 (5) |
| Low Physical Activity (%) | 19 | 16 | 14 | 17 |
| Health Eating Score<sup>b</sup> ($\bar{x}$, $sd$) | 31 (6) | 37 (6) | 36 (7) | 36 (7) |
| Baseline chronic condition (%) | 15 | 21 | 17 | 16 |
| Medications for CVD (%) | 8 | 14 | 9 | 11 |
| Hypertension (%) | 39 | 41 | 40 | 42 |
| Outdoor PM<sub>2.5</sub> ($\mu\text{g}/\text{m}^3$) ($\bar{x}$, $sd$) | 25 (13) | 54.5 (29) | 21 (18) | 41.3 (23) |
| Individuals by Country (%) |  |  |  |  |
| China | 5 | 57 | 22 | 52 |
| India | 50 | 38 | 18 | 36 |
| Africa (South Africa & Tanzania) | 45 | 5 | 60 | 12 |

<sup>a</sup> INTERHEART risk score is a composite measure of multiple modifiable risk factors for MI (Higher values indicate larger risk).
<sup>b</sup> Healthy Eating Score is a composite measure of diet (Higher values indicate a healthier diet).

However, kerosene users also had lower INTERHEART risk scores and lower CVD medication use.

### Baseline cardiorespiratory symptoms

Cardiorespiratory symptoms at baseline were significantly more common amongst kerosene compared to clean fuel users (Table 2). For example, kerosene use was associated with an adjusted Odds Ratio (OR) of 1.44 (95% CI: 1.21-1.72) for chronic cough or sputum; 1.36 (95% CI: 1.17-1.57) for chest pain or tightness with usual activity; and 1.34 (95% CI: 1.15, 1.57) for dyspnea with usual activity.

**Table 2.**
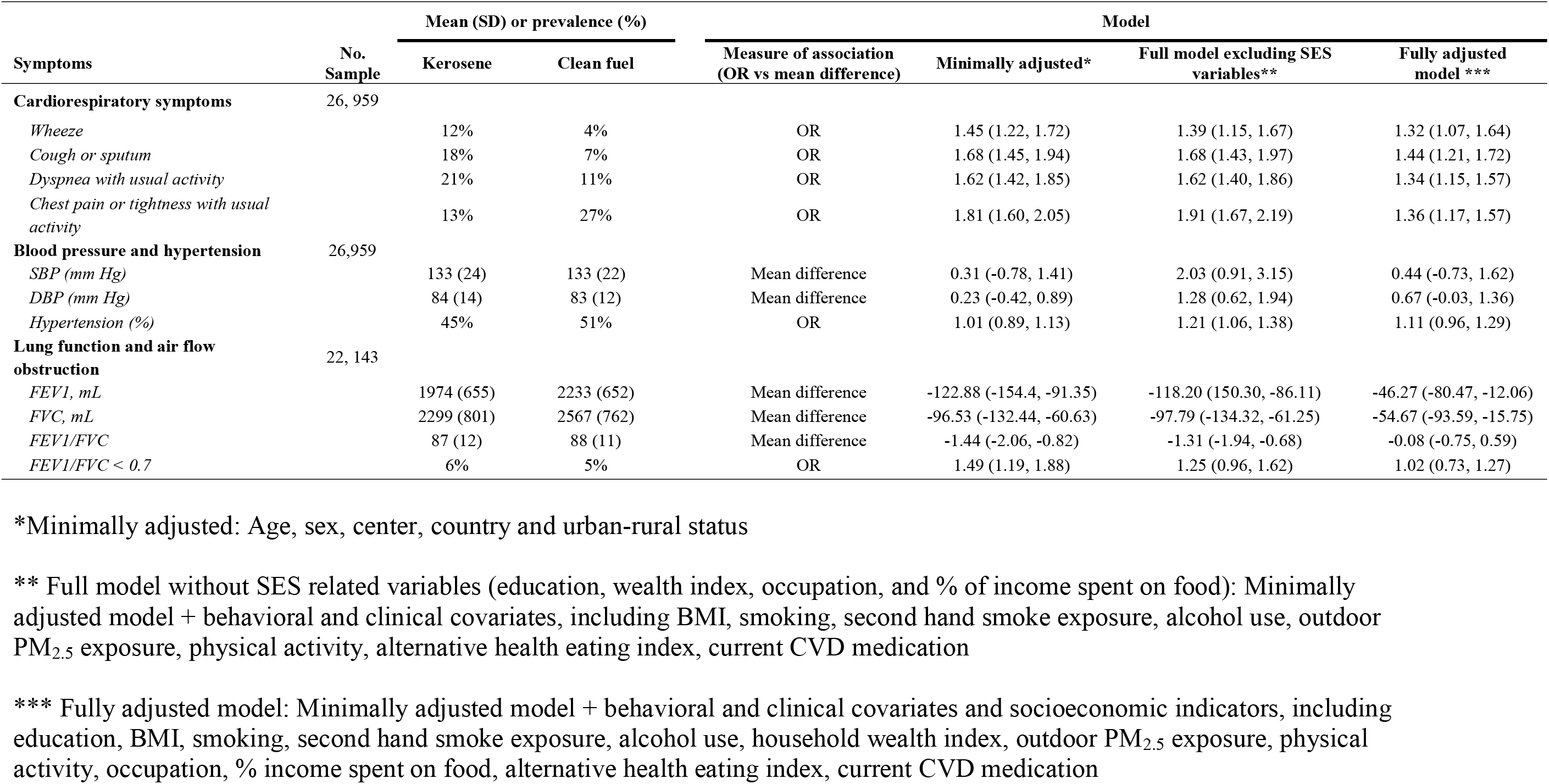
**Summary, odds ratio, and mean difference for symptoms, BP, and lung function among those using kerosene compared to those using clean cooking fuels**

| Symptoms | No. Sample | Mean (SD) or prevalence (%) |  | Measure of association (OR vs mean difference) | Model |  |  |
| --- | --- | --- | --- | --- | --- | --- | --- |
|  |  | Kerosene | Clean fuel |  | Minimally adjusted* | Full model excluding SES variables** | Fully adjusted model *** |
| **Cardiorespiratory symptoms** | 26, 959 |  |  |  |  |  |  |
| Wheeze |  | 12% | 4% | OR | 1.45 (1.22, 1.72) | 1.39 (1.15, 1.67) | 1.32 (1.07, 1.64) |
| Cough or sputum |  | 18% | 7% | OR | 1.68 (1.45, 1.94) | 1.68 (1.43, 1.97) | 1.44 (1.21, 1.72) |
| Dyspnea with usual activity |  | 21% | 11% | OR | 1.62 (1.42, 1.85) | 1.62 (1.40, 1.86) | 1.34 (1.15, 1.57) |
| Chest pain or tightness with usual activity |  | 13% | 27% | OR | 1.81 (1.60, 2.05) | 1.91 (1.67, 2.19) | 1.36 (1.17, 1.57) |
| **Blood pressure and hypertension** | 26,959 |  |  |  |  |  |  |
| SBP (mm Hg) |  | 133 (24) | 133 (22) | Mean difference | 0.31 (-0.78, 1.41) | 2.03 (0.91, 3.15) | 0.44 (-0.73, 1.62) |
| DBP (mm Hg) |  | 84 (14) | 83 (12) | Mean difference | 0.23 (-0.42, 0.89) | 1.28 (0.62, 1.94) | 0.67 (-0.03, 1.36) |
| Hypertension (%) |  | 45% | 51% | OR | 1.01 (0.89, 1.13) | 1.21 (1.06, 1.38) | 1.11 (0.96, 1.29) |
| **Lung function and air flow obstruction** | 22, 143 |  |  |  |  |  |  |
| FEV1, mL |  | 1974 (655) | 2233 (652) | Mean difference | -122.88 (-154.4, -91.35) | -118.20 (150.30, -86.11) | -46.27 (-80.47, -12.06) |
| FVC, mL |  | 2299 (801) | 2567 (762) | Mean difference | -96.53 (-132.44, -60.63) | -97.79 (-134.32, -61.25) | -54.67 (-93.59, -15.75) |
| FEV1/FVC |  | 87 (12) | 88 (11) | Mean difference | -1.44 (-2.06, -0.82) | -1.31 (-1.94, -0.68) | -0.08 (-0.75, 0.59) |
| FEV1/FVC < 0.7 |  | 6% | 5% | OR | 1.49 (1.19, 1.88) | 1.25 (0.96, 1.62) | 1.02 (0.73, 1.27) |

\*Minimally adjusted: Age, sex, center, country and urban-rural status
\*\* Full model without SES related variables (education, wealth index, occupation, and % of income spent on food): Minimally adjusted model + behavioral and clinical covariates, including BMI, smoking, second hand smoke exposure, alcohol use, outdoor PM<sub>2.5</sub> exposure, physical activity, alternative health eating index, current CVD medication
\*\*\* Fully adjusted model: Minimally adjusted model + behavioral and clinical covariates and socioeconomic indicators, including education, BMI, smoking, second hand smoke exposure, alcohol use, household wealth index, outdoor PM<sub>2.5</sub> exposure, physical activity, occupation, % income spent on food, alternative health eating index, current CVD medication

### Baseline blood pressure and hypertension

Overall, kerosene and clean fuel users had similar mean (standard deviation) systolic and diastolic BP of 133 (22) mmHg and 83 (12) mmHg, respectively (Table 2). Approximately 40% of the participants and those each in kerosene and clean fuel groups had hypertension. After adjustment for demographic, behavioral, clinical, and SES variables, there was no clear difference in BP in those using kerosene compared to those using clean fuels for cooking while the OR for hypertension was somewhat elevated after covariate adjustment (Table 2).

### Baseline lung function and air flow obstruction

Although attenuated with extensive covariate adjustment, we found lower lung function in kerosene users compared to clean fuel users by 46 and 55 mL, for FEV1 and FVC, respectively (Table 2). Similarly, the adjusted cross-sectional age-related mean differences in FEV1/FVC ratio were lower in kerosene users compared to clean fuel users (Figure 2). Based upon FEV1/FVC < 0.7, we observed that 8% of kerosene users had air flow obstruction versus 6% of the clean fuel group at baseline. The odds of having air flow obstruction among kerosene users relative to clean fuels was 1.49 (95% CI: 1.19, 1.88) in the minimally adjusted model, which was substantially attenuated to 1.02 (95% CI: 0.73, 1.27) in the fully adjusted model (Table 2).

**Figure 2.**
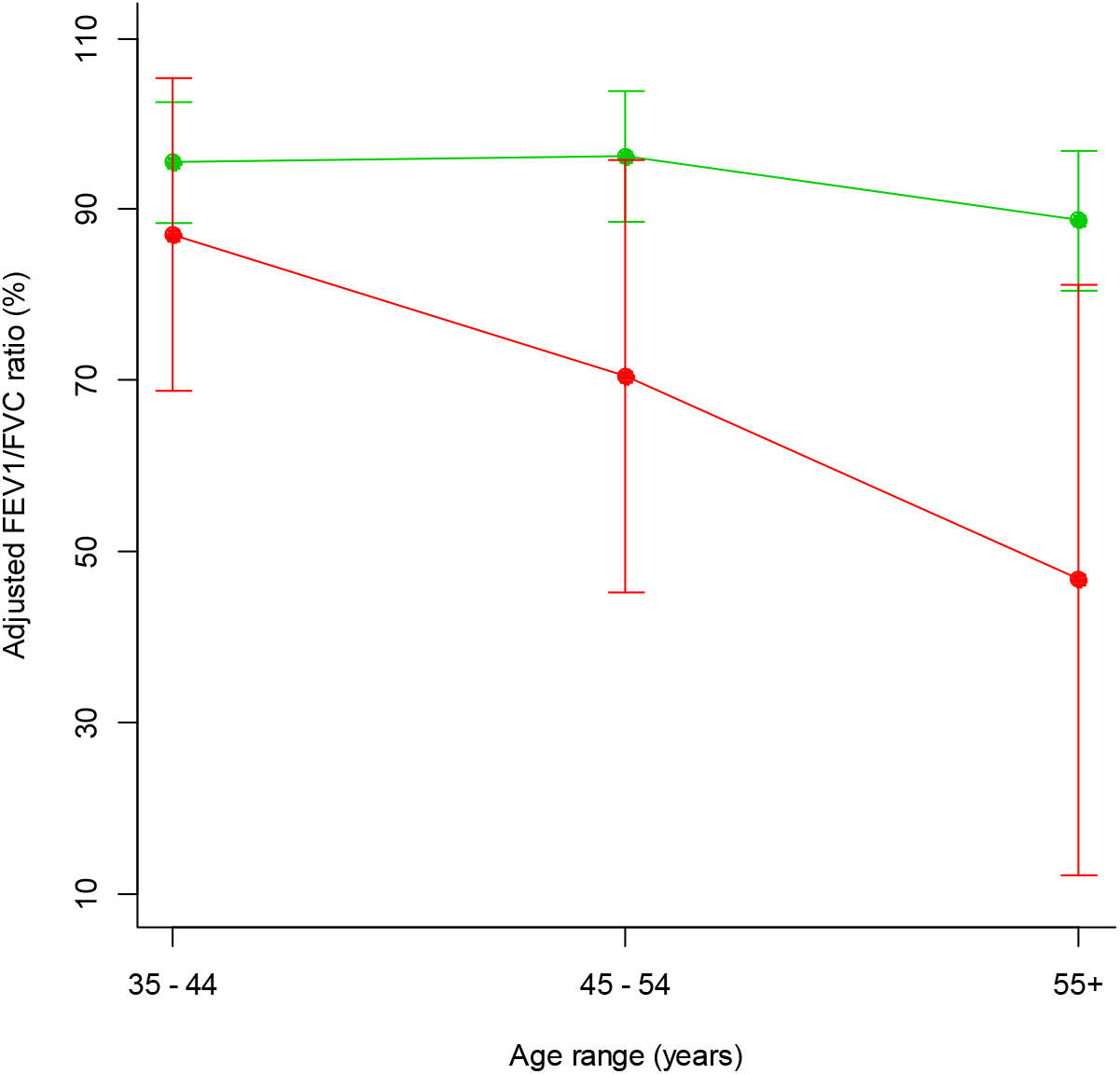
**Adjusted mean FEV1/FVC ratio by age and fuel groups**

Multilevel marginal linear regression was used to model FEV1/FVC ratio by age and fuel groups (kerosene and clean fuel), with sex, BMI, education, ethnicity, smoking, and rural/urban location as covariates, and random effects for country and center.

### Follow-up events

We recorded 1,894 deaths, 1,708 major CVD events, and 353 major respiratory diseases during follow-up (median of 9.4 years). In the fully adjusted model, all-cause mortality, major CVD, and respiratory disease were higher in households cooking primarily with kerosene compared to homes primarily using clean fuels [HR = 1.32 (95% CI: 1.14, 1.53), HR = 1.34 (95% CI: 1.08, 1.66), and HR= 1.55 (95% CI: 0.98, 2.43) respectively] (Table 3). The composite measure, which combined all of the first major CVD or respiratory event or death, also demonstrated increased risk with primary use of kerosene compared to clean fuels (HR = 1.36 [95% CI: 1.19, 1.56]).

**Table 3.**
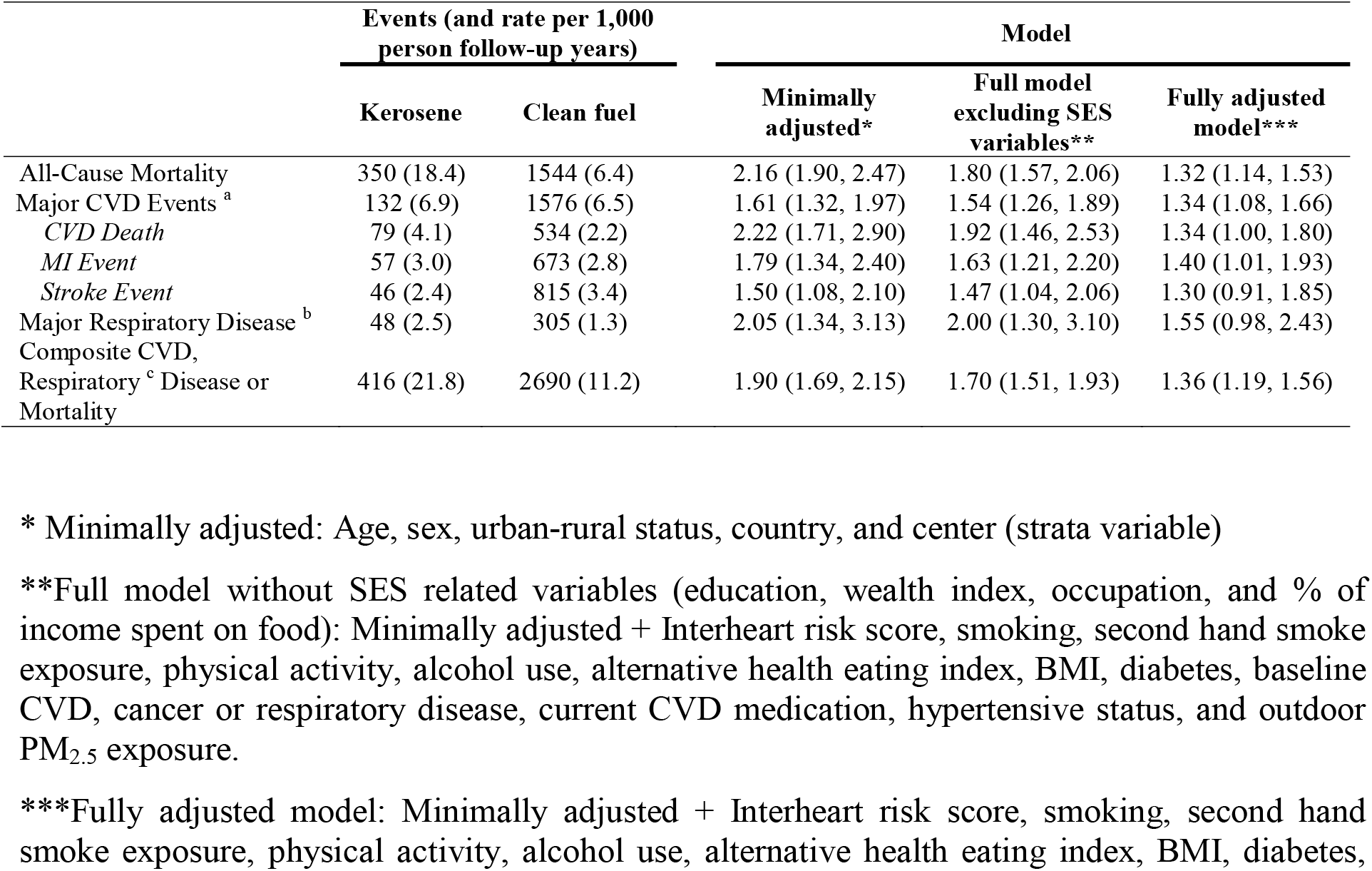
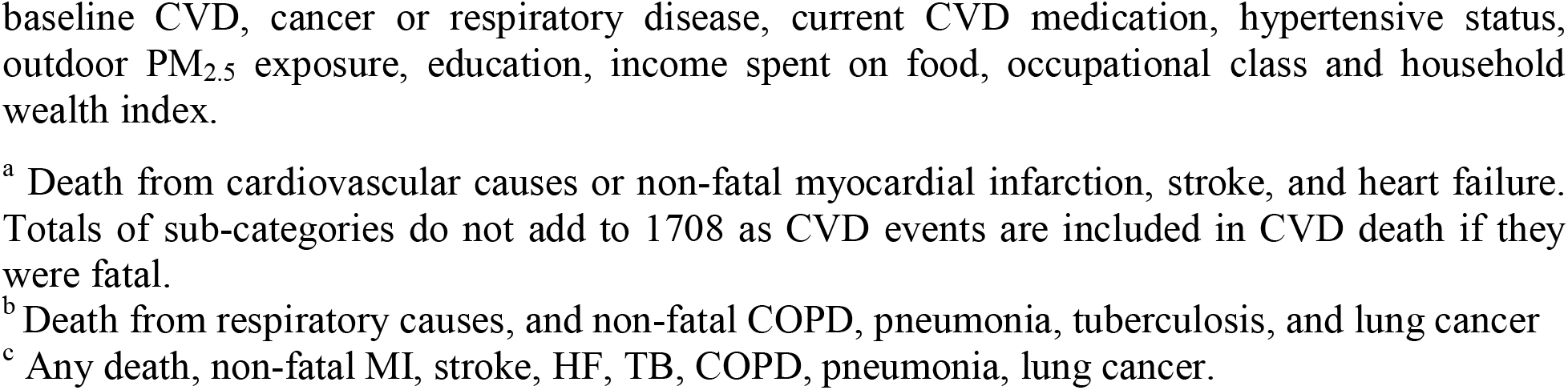
**Hazard ratios for the associations between kerosene use for cooking, compared to clean fuels, with all-cause mortality, CVD and respiratory diseases**

### Effect of kerosene use primarily for cooking relative to solid fuel

Although the literature on household air pollution is dominated by comparing solid to clean fuels, sociodemographic factors are generally more similar among kerosene and solid fuel users than for those using clean fuels. Accordingly, we also compared the effect of kerosene to solid fuel use for cooking for the outcomes examined above (Table 4). Compared to primary use of solid fuels, we observed elevated risks for primary use of kerosene in both minimally and fully adjusted models for all outcomes. For instance, in the full model, we found between 33-49% higher ORs for cardiorespiratory symptoms, 20-68% higher for all-cause mortality and CVD events, and about 20% higher for hypertension in kerosene users compared to solid fuel users (Table 4). While we still observed substantial attenuation of effect estimates following covariate adjustment, the overall results indicate a larger risk of respiratory morbidity and cardiorespiratory mortality for those using kerosene as a primary cooking fuel compared to those cooking primarily with solid fuels (Table 4).

**Table 4.**
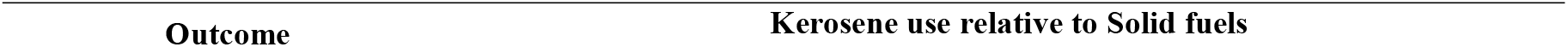
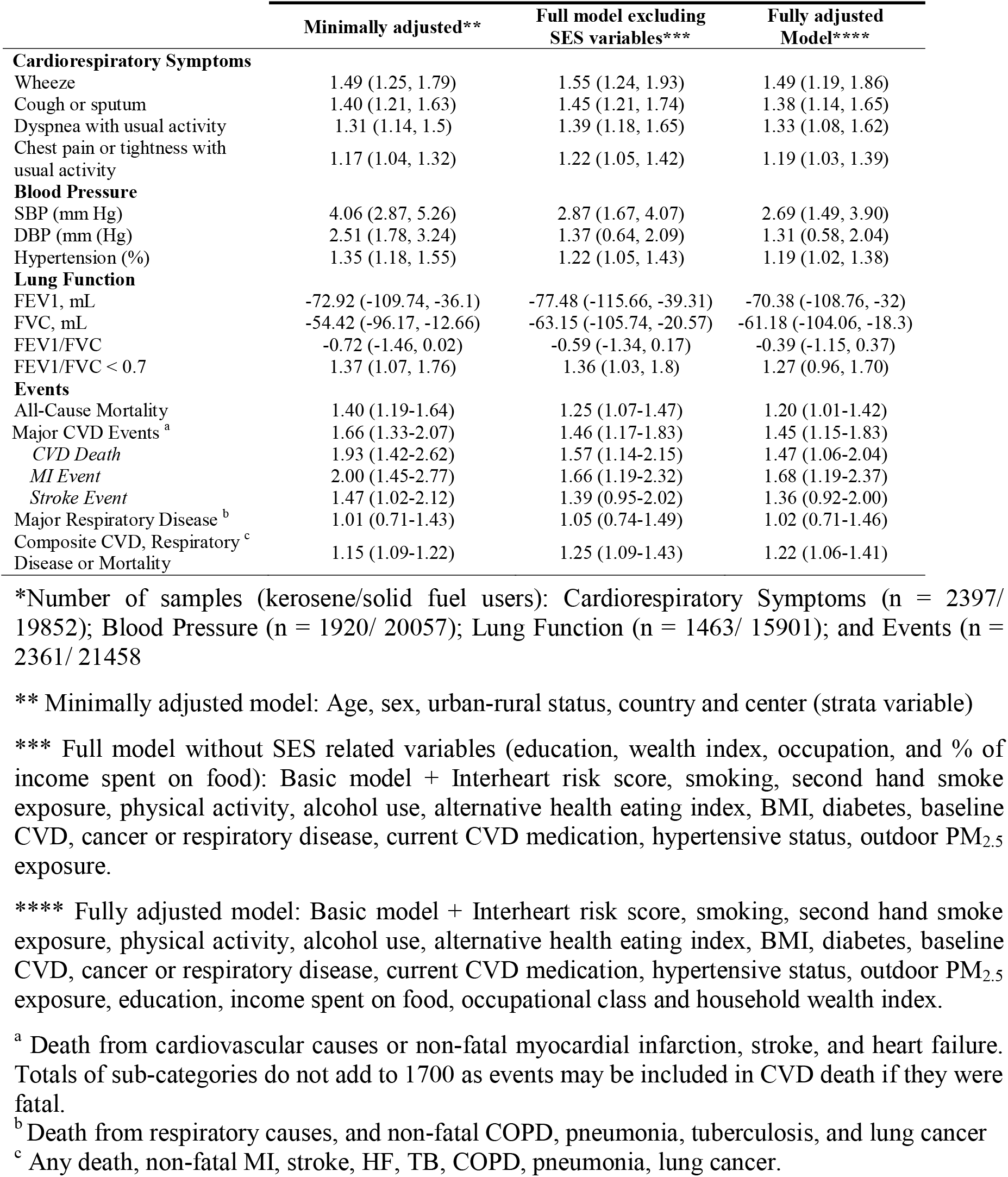
**Estimates comparing primary use of kerosene for cooking to primary use of solid fuel users for all outcomes\***

| Outcome | Kerosene use relative to Solid fuels |
| --- | --- |

|  | Minimally adjusted** | Full model excluding SES variables*** | Fully adjusted Model**** |
| --- | --- | --- | --- |
| **Cardiorespiratory Symptoms** |  |  |  |
| Wheeze | 1.49 (1.25, 1.79) | 1.55 (1.24, 1.93) | 1.49 (1.19, 1.86) |
| Cough or sputum | 1.40 (1.21, 1.63) | 1.45 (1.21, 1.74) | 1.38 (1.14, 1.65) |
| Dyspnea with usual activity | 1.31 (1.14, 1.5) | 1.39 (1.18, 1.65) | 1.33 (1.08, 1.62) |
| Chest pain or tightness with usual activity | 1.17 (1.04, 1.32) | 1.22 (1.05, 1.42) | 1.19 (1.03, 1.39) |
| **Blood Pressure** |  |  |  |
| SBP (mm Hg) | 4.06 (2.87, 5.26) | 2.87 (1.67, 4.07) | 2.69 (1.49, 3.90) |
| DBP (mm (Hg) | 2.51 (1.78, 3.24) | 1.37 (0.64, 2.09) | 1.31 (0.58, 2.04) |
| Hypertension (%) | 1.35 (1.18, 1.55) | 1.22 (1.05, 1.43) | 1.19 (1.02, 1.38) |
| **Lung Function** |  |  |  |
| FEV1, mL | -72.92 (-109.74, -36.1) | -77.48 (-115.66, -39.31) | -70.38 (-108.76, -32) |
| FVC, mL | -54.42 (-96.17, -12.66) | -63.15 (-105.74, -20.57) | -61.18 (-104.06, -18.3) |
| FEV1/FVC | -0.72 (-1.46, 0.02) | -0.59 (-1.34, 0.17) | -0.39 (-1.15, 0.37) |
| FEV1/FVC < 0.7 | 1.37 (1.07, 1.76) | 1.36 (1.03, 1.8) | 1.27 (0.96, 1.70) |
| **Events** |  |  |  |
| All-Cause Mortality | 1.40 (1.19-1.64) | 1.25 (1.07-1.47) | 1.20 (1.01-1.42) |
| Major CVD Events <sup>a</sup> | 1.66 (1.33-2.07) | 1.46 (1.17-1.83) | 1.45 (1.15-1.83) |
| CVD Death | 1.93 (1.42-2.62) | 1.57 (1.14-2.15) | 1.47 (1.06-2.04) |
| MI Event | 2.00 (1.45-2.77) | 1.66 (1.19-2.32) | 1.68 (1.19-2.37) |
| Stroke Event | 1.47 (1.02-2.12) | 1.39 (0.95-2.02) | 1.36 (0.92-2.00) |
| Major Respiratory Disease <sup>b</sup> | 1.01 (0.71-1.43) | 1.05 (0.74-1.49) | 1.02 (0.71-1.46) |
| Composite CVD, Respiratory <sup>c</sup> Disease or Mortality | 1.15 (1.09-1.22) | 1.25 (1.09-1.43) | 1.22 (1.06-1.41) |

\*Number of samples (kerosene/solid fuel users): Cardiorespiratory Symptoms (n = 2397/ 19852); Blood Pressure (n = 1920/ 20057); Lung Function (n = 1463/ 15901); and Events (n = 2361/ 21458)
\*\* Minimally adjusted model: Age, sex, urban-rural status, country and center (strata variable)
\*\*\* Full model without SES related variables (education, wealth index, occupation, and % of income spent on food): Basic model + Interheart risk score, smoking, second hand smoke exposure, physical activity, alcohol use, alternative health eating index, BMI, diabetes, baseline CVD, cancer or respiratory disease, current CVD medication, hypertensive status, outdoor PM<sub>2.5</sub> exposure.
\*\*\*\* Fully adjusted model: Basic model + Interheart risk score, smoking, second hand smoke exposure, physical activity, alcohol use, alternative health eating index, BMI, diabetes, baseline CVD, cancer or respiratory disease, current CVD medication, hypertensive status, outdoor PM<sub>2.5</sub> exposure, education, income spent on food, occupational class and household wealth index.
<sup>a</sup> Death from cardiovascular causes or non-fatal myocardial infarction, stroke, and heart failure. Totals of sub-categories do not add to 1700 as events may be included in CVD death if they were fatal.
<sup>b</sup> Death from respiratory causes, and non-fatal COPD, pneumonia, tuberculosis, and lung cancer
<sup>c</sup> Any death, non-fatal MI, stroke, HF, TB, COPD, pneumonia, lung cancer.

### Sensitivity analyses

In sensitivity analysis, we observed large differences in sociodemographic measures between kerosene and clean fuel users that led to substantial attenuation of effect estimates between minimally and fully adjusted models. We examined the potential for residual confounding by comparing the models with or without key variables related to SES and observed, in general, 10-60% reductions in the point estimates when the SES variables were added to the model including individual and household risk factors (Table 2 and 3). Further, we stratified the models by the same SES indicators and observed similar effect estimates across strata of education for the follow-up events and mortality (Table 5). Inconsistent stratum specific estimates for other outcomes by education, household wealth, and percent of income spent on food (Smith and Wiesmann 2007) suggest some potential for residual confounding in our models. Households with low food security were mostly in Tanzania and South Africa, where we also observed weak effect sizes across several outcomes. When models were restricted to India and China and to households spending <50% of their income on food, we observed similar associations with mortality (HR: 1.63, 95% CI: 1.34-1.97) and major CVD (HR: 1.58, 95% CI: 1.22-2.05) as our main result. The effect of occupational class was mixed, as we observed inconsistent relationships for skilled/professional workers compared to unskilled workers. Surprisingly, we generally observed weak associations for homemakers, where we would expect higher exposure levels to cooking with kerosene.

**Table 5.** **Adjusted models* comparing kerosene with clean fuel for both baseline and follow-up health measures stratified by key socio-economic variables**

|  | Dyspnea with usual activity | Chest pain or tightness with usual activity | FEV1/FVC <0.7 | Hypertension | SBP | DBP | FEV1 | All-Cause Mortality | Major CVD Events<sup>a</sup> | Composite CVD, Respiratory Event or Mortality |
| --- | --- | --- | --- | --- | --- | --- | --- | --- | --- | --- |
|  | OR (95% CI) |  |  |  | Mean Difference (95% CI) |  |  | HR (95% CI) |  |  |
| **Education** |  |  |  |  |  |  |  |  |  |  |
| ≤ Primary | 1.30 (1.06, 1.58) | 1.31 (1.09, 1.58) | 1.05 (0.75, 1.47) | 1.02 (0.85, 1.24) | -0.50 (-2.28, 1.28) | 0.50 (-0.51, 1.51) | 1.83 (-45.98, 49.65) | 1.33 (1.10-1.60) | 1.28 (0.97-1.69) | 1.30 (1.10-1.54) |
| > Primary | 1.18 (0.89, 1.56) | 1.28 (0.99, 1.65) | 0.68 (0.39, 1.16) | 1.14 (0.89, 1.44) | 1.80 (0.00, 3.60) | 0.79 (-0.28, 1.87) | -33.92 (-86.25, 18.41) | 1.32 (1.01-1.71) | 1.39 (0.97-2.00) | 1.44 (1.15-1.80) |
| **Wealth Index** |  |  |  |  |  |  |  |  |  |  |
| Lowest (T1) | 1.25 (0.96, 1.64) | 1.03 (0.80, 1.33) | 1.11 (0.72, 1.73) | 0.95 (0.74, 1.23) | 1.16 (-1.13, 3.45) | 0.27 (-1.03, 1.58) | -5.57 (-75.32, 64.18) | 1.38 (1.05-1.82) | 1.70 (1.16-2.50) | 1.46 (1.15-1.86) |
| Moderate (T2) | 1.31 (1.03, 1.66) | 1.46 (1.17, 1.81) | 0.96 (0.61, 1.52) | 1.13 (0.89, 1.42) | -0.31 (-2.13, 1.52) | 0.38 (-0.7, 1.46) | -81.51 (-133.68, -29.34) | 1.36 (1.09-1.69) | 1.35 (0.97-1.88) | 1.31 (1.07-1.60) |
| Highest (T3) | 1.08 (0.66, 1.78) | 1.09 (0.69, 1.71) | 0.95 (0.41, 2.23) | 1.34 (0.88, 2.02) | -0.61 (-3.89, 2.68) | 0.12 (-1.82, 2.05) | -57.66 (-144.63, 29.31) | 1.35 (0.89-2.07) | 1.32 (0.71-2.43) | 1.42 (0.97-2.07) |
| **Occupation** |  |  |  |  |  |  |  |  |  |  |
| Skilled/ Professional | 1.24 (0.96, 1.60) | 1.46 (1.16, 1.83) | 0.98 (0.53, 1.83) | 1.08 (0.85, 1.37) | 0.97 (-0.85, 2.79) | 1.26 (0.19, 2.33) | -66.86 (-123.01, -10.71) | 1.69 (0.95-3.02) | 1.59 (0.81-3.14) | 1.60 (1.01-2.53) |
| Unskilled workers | 1.49 (1.00, 2.23) | 1.17 (0.8, 1.7) | 1.1 (0.63, 1.92) | 1.19 (0.86, 1.65) | 0.85 (-2.01, 3.7) | 0.63 (-1.1, 2.36) | -5.14 (-93.2, 82.91) | 1.22 (0.88-1.70) | 1.05 (0.66-1.69) | 1.37 (1.03-1.83) |
| Homemaker | 0.97 (0.75, 1.26) | 0.83 (0.65, 1.07) | 0.89 (0.59, 1.34) | 0.92 (0.72, 1.18) | -0.89 (-3.1, 1.32) | -1.20 (-2.47, 0.07) | 0.35 (-52.57, 53.27) | 1.05 (0.83-1.34) | 1.09 (0.75-1.57) | 1.05 (0.84-1.31) |
| **% Income Spent on Food** |  |  |  |  |  |  |  |  |  |  |
| ≥ 50% | 1.22 (0.99, 1.49) | 1.33 (1.11, 1.61) | 1.00 (0.66, 1.52) | 1.13 (0.93, 1.38) | 1.62 (0.02, 3.23) | 1.01 (0.07, 1.95) | -88.58 (-133.53, -43.62) | 1.38 (1.14-1.68) | 1.24 (0.92-1.67) | 1.35 (1.13-1.62) |
| < 50% | 1.39 (1.04, 1.86) | 1.21 (0.92, 1.59) | 0.88 (0.58, 1.34) | 0.96 (0.74, 1.23) | -0.29 (-2.31, 1.73) | 0.37 (-0.82, 1.57) | 7.26 (-50.03, 64.55) | 0.99 (0.76-1.29) | 1.18 (0.83-1.68) | 1.06 (0.84-1.35) |
| **Gender** |  |  |  |  |  |  |  |  |  |  |
| Female | 1.33 (1.1, 1.61) | 1.43 (1.19, 1.7) | 1.04 (0.72, 1.50) | 1.17 (0.98, 1.4) | 1.21 (-0.33, 2.74) | 1.00 (0.13, 1.87) | -7.78 (-45.43, 29.88) | 1.25 (1.01, 1.54) | 1.30 (0.96, 1.77) | 1.31 (1.08, 1.58) |
| Male | 1.37 (1.05, 1.8) | 1.27 (0.99, 1.63) | 0.93 (0.62, 1.40) | 0.94 (0.75, 1.18) | -0.65 (-2.48, 1.18) | 0.11 (-1.03, 1.25) | -85.52 (-145.99, -25.04) | 1.35 (1.10, 1.66) | 1.44 (1.06, 1.95) | 1.40 (1.16, 1.70) |
| **Urban/Rural** |  |  |  |  |  |  |  |  |  |  |
| Urban | 1.15 (0.94, 1.39) | 1.26 (1.05, 1.5) | 1.00 (0.7, 1.43) | 1.05 (0.88, 1.26) | -0.84 (-2.26, 0.58) | 0.41 (-0.42, 1.25) | -85.55 (-126.12, -44.99) | 1.24 (1.02-1.51) | 1.02 (0.76-1.36) | 1.21 (1.01-1.44) |
| Rural | 1.42 (1.06, 1.91) | 1.16 (0.87, 1.56) | 0.97 (0.58, 1.60) | 0.98 (0.74, 1.29) | -0.18 (-2.51, 2.14) | -0.17 (-1.54, 1.20) | 1.36 (-65.37, 68.1) | 1.04 (0.77-1.39) | 1.43 (0.96-2.11) | 1.16 (0.90-1.49) |
| **Country** |  |  |  |  |  |  |  |  |  |  |
| India | 1.35 (1.09, 1.67) | 1.42 (1.17, 1.72) | 1.08 (0.63, 1.84) | 1.16 (0.95, 1.43) | 1.25 (-0.34, 2.84) | 1.52 (0.58, 2.45) | -127.27 (-171.33, -83.2) | 1.44 (1.17-1.77) | 1.31 (0.96-1.79) | 1.43 (1.18-1.73) |
| China | 1.56 (0.95, 2.57) | 1.27 (0.78, 2.07) | 0.77 (0.42, 1.41) | 0.80 (0.55, 1.17) | -1.23 (-4.31, 1.86) | -1.21 (-3.01, 0.60) | 94.64 (9.59, 179.68) | 0.71 (0.34-1.48) | 1.49 (0.93-2.40) | 1.38 (0.92-2.07) |
| Africa (Tanzania and South Africa) | 0.86 (0.63, 1.15) | 0.71 (0.54, 0.95) | 0.72 (0.45, 1.14) | 0.93 (0.69, 1.25) | -2.21 (-5.00, 0.59) | -1.37 (-3.06, 0.33) | 17.93 (-48.56, 84.43) | 1.03 (0.79-1.35) | 1.08 (0.68-1.70) | 1.05 (0.82-1.35) |

\*Full model: Age, sex, urban-rural status, country, center (strata variable), interheart risk score, smoking, second hand smoke exposure, physical activity, alcohol use, alternative health eating index, BMI, diabetes, baseline CVD, cancer or respiratory disease, current CVD medication, hypertensive status, outdoor PM<sub>2.5</sub> exposure, education, income spent on food, occupational class and household wealth index

## Discussion

In its 2014 indoor air quality guidelines, the World Health Organization (WHO) discouraged household use of kerosene until further research into its health impacts became available (World Health Organization 2014). Since then however, no prospective cohort studies assessing cardiorespiratory risks associated with household kerosene use have been published. Lack of prospective studies hinder assessment of the impact of kerosene use on incident disease. To our knowledge, this is the first prospective study to demonstrate the adverse impacts of kerosene use for cooking on mortality and cardiorespiratory health outcomes. In examining multiple but related outcomes spanning from clinical measures to mortality at baseline and prospectively, we demonstrated consistency and coherence in our results. We used follow-up data on cardiorespiratory events across multiple countries and communities and found increased risks for all-cause mortality, major CVD, and major respiratory disease for individuals that cook primarily with kerosene, compared to those cooking with clean fuels. We also observed consistently higher odds of baseline respiratory symptoms, and impaired lung function amongst kerosene users compared with those who cooked primarily with clean fuels. The relative decrease in lung function in kerosene users was especially pronounced in older age groups. Additionally, we observed increased risks for kerosene use, compared to solid fuel use for all outcomes, except airflow limitation (FEV_1_/FVC). These results suggest that cooking with kerosene may well be associated with worse cardiorespiratory health than the use of solid fuels.

The limited measurements of emissions from kerosene cook stoves found high levels of gaseous pollutants (e.g. carbon monoxide and Sulphur dioxide) and PM concentrations, which are considered a robust indicator of health-relevant pollutant mixtures arising from cooking (Lam et al. 2012b). Our findings and the biological plausibility of the observed associations between kerosene use and cardiovascular and respiratory health impacts are supported by experimental evidence indicating the ability of kerosene combustion to induce inflammation and production of reactive oxygen species as well as degrade the effectiveness of pulmonary surfactants (Lam et al. 2012b; Maiyoh et al. 2015). However, while kerosene use is still common in many LMICs, there has been limited epidemiological studies on its use for cooking, especially from longitudinal studies. Our analysis using a median of 9.4 years of follow-up data demonstrated increased risks for all-cause mortality (32%, 95% CI: 14 – 53%), CVD events (34%, 95% CI: 8 – 66%), and respiratory disease (55%, 95% CI: -2 – 1.43%) for individuals cooking with kerosene. Our findings of higher risk of death among kerosene users supports and extends the work of Mitter and colleagues (Mitter et al. 2016), which reported a HR of 6% (95% CI: 2 – 10%) for all-cause mortality for kerosene/diesel burning relative to wood, dung, and gas in a prospective cohort in Iran. We are not aware of any additional prospective studies examining kerosene use for cooking with mortality and major CVD and respiratory disease.

Since the existing literature is focused mainly on comparisons between clean and solid fuels, we compare our results broadly with other sources of household air pollution to show that our findings were consistent with studies that evaluated primary use of solid fuels for cooking versus clean fuels. For example, the China Kadoorie Biobank study of 271,217 participants observed that the use of solid fuels for cooking was associated with a HR of 1.11 (95% CI: 1.03-1.20) and 1.20 (95% CI: 1.02-1.41) for all-cause and cardiovascular mortality, respectively (Yu et al. 2018). A retrospective cohort study of 22,337 individuals 18 years of age or older in rural Bangladesh found incident rate ratios of 2.26 (95% CI: 1.02-4.99) for respiratory mortality, 1.10 (0.89-1.37) for non-communicable disease mortality and 1.07 (95% CI: 0.82-1.41) for CVD mortality when comparing solid fuel use for cooking to gas (Alam et al. 2012). Finally, for 74,941 women followed in Shanghai, those who used coal for cooking, compared to those who never used coal, had increased risks for all-cause mortality (1.12, 95% CI: 1.05-1.21), and CVD mortality [1.18 (95% CI: 1.02-1.37)] (Kim et al. 2016). In the PURE cohort itself, compared to electricity or gas, living in households with solid fuel use (n=88,431 adults) was associated with increases of 1.12 (95% CI: 1.04-1.21) for all-cause mortality, 1.08 (95% CI: 0.99-1.17) for CVD, and 1.14 (95% CI: 1.00-1.30) for respiratory disease (Hystad et al. 2019). A composite measure of all the outcomes examined in Hystad and colleagues indicated a HR of 1.12 (95% CI: 1.06-1.19) for solid fuel compared with clean fuel use (Hystad et al. 2019). More importantly, while previous studies have largely focused on solid fuels against clean fuels, we have estimated higher risks with kerosene than with solid fuels, indicating that kerosene use may lead to worse cardiorespiratory health than use of solid fuels.

Our cross-sectional analyses of baseline measurements also indicate that kerosene use was associated with higher prevalence rate of respiratory symptoms and with lower lung function. Previous studies reporting such links have been small in sample size. In line with our baseline findings, Choi et al (Choi et al. 2015) reported in a cross-sectional study, higher rates of self-reported cough, phlegm, and chest pain in 547 women in Bangladesh who used kerosene versus LPG. Our observations of reduced lung function, together with higher prevalence of respiratory symptoms indicate higher rates of likely undiagnosed chronic obstructive pulmonary disease in kerosene users compared with clean fuel users. Past studies that have evaluated spirometry records with kerosene use were mostly conducted in children (Lam et al. 2012a). In relation to solid fuels, our cross-sectional analyses indicate higher risks from kerosene use than solid fuels on baseline cardiorespiratory symptoms.

With a relatively large sample size from a multi-center/multi-country cohort, and detailed data on SES indicators and other characteristics, we were able to adjust for many potentially important confounders in both cross-sectional and longitudinal analyses. The increased risk of cardiorespiratory symptoms and cardiovascular events implicate kerosene use as a risk factor for cardiovascular morbidity and mortality. Our results further support the transition towards clean fuel use, with the recommendation by the WHO discouraging household use of kerosene (World Health Organization 2014). Our overall findings provide evidence that kerosene is an unhealthy household fuel and suggest that the contribution of kerosene to the burden of household air pollution-related disease and deaths in LMICs has been under-recognized. Our findings therefore support the recommendation by the international global health community to no longer consider kerosene as ‘clean’.

## Strengths and limitations

While the PURE study has several benefits, such as a large sample size from an international prospective cohort and standardized approaches for data collection and outcome ascertainment, there are some important limitations to this analysis. Kerosene use was associated with much lower individual and household SES. Kerosene use is closely tied to poverty, and while we were able to adjust our models for many potential confounders, including several poverty-related factors (such as low income, low education and rural location), we cannot fully rule out residual confounding from potential unknown, unmeasured, or imperfectly measured factors. However, it is unlikely that such residual confounding explains our results. We calculated e-values for the observed HR associations (a measure of the minimum strength required for both the exposure-confounder and exposure-disease relationships to ‘explain away’ the estimated relationship between exposure and disease (VanderWeele and Ding 2017)). For all-cause mortality, the observed HR was 1.32 compared to an e-value of 1.97, suggesting an unmeasured confounder needs to be associated with both kerosene use and all-cause mortality by a risk ratio of 1.97-fold each, above and beyond the measured confounders included in the model. We observed much smaller reductions in the point estimates (10-60%) when all SES variables were added to a model with individual and household risk factors. It is therefore unlikely that there would be residual poverty related effects or unmeasured factors that are completely unrelated to the comprehensive individual, household and community variables included in our models. Stratified analyses also showed consistent associations within lower SES strata, suggesting minimal potential for residual confounding. In addition, our ability to assess symptoms at baseline and related outcomes in follow-up data, as well as the coherent findings across multiple cardiorespiratory outcomes adds to the validity of our findings.

Another limitation of our study is that we relied on a single measure of primary fuel use for cooking to assess exposure (kerosene use vs gas or electricity or solid fuels). While the use of household’s primary cooking fuel type as a single indicator of household air pollution exposure is commonplace in large population-based studies, this likely introduced exposure misclassification due to factors like the use of kerosene for lighting, stove stacking, ventilation, cooking practices and user behavior patterns, housing characteristics and emissions from other sources (Bruce et al. 2015; Clark et al. 2013; Smith et al. 2014). Although technically possible, it would be too costly and logistically prohibitive in terms of budget, time, resources and participant burden to measure detailed personal exposures in such a large multi-country/center study designs as ours, especially given how interrelated household and ambient exposures are in LMIC settings where biomass fuel use is common. Additionally, we only collected data from participants on cooking fuels and have no information on other uses of kerosene such as lighting. Fuel use was also self-reported, which could suffer from reliability and validity of the reported exposure. However, because we used a standardized protocol to collect data across study centers, biases from self-reported fuel use would likely equally affect all study centers and across both the exposed versus unexposed groups. Further, we did not have data on household fuel switching during follow-up, which was likely in some households (Shupler et al. 2019). Finally, while event identification relied on a number of steps such as face to face interviews, certified hospital records, death certificates, and verbal autopsies, our data are subject to some recall bias which could have misclassified some events. Nevertheless, the final event data used in these analyses were adjudicated in each county by trained physicians following a standardized procedure to improve data reliability.

## Conclusion

While the WHO discourages the use of kerosene in households because of the potential health risks, kerosene is still widely used in developing countries for cooking, heating and lighting. For policy and practice, our absolute (compared with clean fuels) and relative (compared to solid fuels) comparisons in this cohort provide evidence that household use of kerosene for cooking increases the risk of cardiovascular and respiratory mortality. Our data also suggested that household use of kerosene for cooking may be more harmful to health than use of solid fuels. Kerosene should be considered an important risk factor for chronic disease and be replaced with cleaner alternatives.

## Data Availability

Request for data can be made to the PURE Study center at http://www.phri.ca

## Funding

This work was supported by the Office Of The Director, National Institutes Of Health of the National Institutes of Health [Award Number DP5OD019850]. The content is solely the responsibility of the authors and does not necessarily represent the official views of the National Institutes of Health; and CIHR [grant #136893].

## Notes

### Competing Interest Statement

The authors have declared no competing interest.

